# From Exercise to Strain: Rapid and Accurate Prediction of Femoral Neck Loading

**DOI:** 10.1101/2024.10.20.24315745

**Authors:** Zainab Altai, Andrew T.M. Phillips, Jason Moran, Xiaojun Zhai, Qichang Mei, Bernard X.W. Liew

**Author notes:** **Correspondence:** Zainab Altai, School of Sport, Rehabilitation and Exercise Sciences, University of Essex, Wivenhoe Park, CO4 3SQ, Colchester, Essex, UK.

## Abstract

Femoral neck fractures pose significant morbidity and mortality risks, particularly among osteoporotic patients. This study aims to identify effective exercises for enhancing bone health and develop a neural network model to predict femoral neck strains during exercise using inertial measurement unit (IMU) data. We employed musculoskeletal modeling (MSK) and finite element (FE) analysis to assess femoral neck strains during various ballistic exercises—walking, running, countermovement jumps, squat jumps, unilateral hopping, and bilateral hopping—across three intensity levels: high, moderate, and low. Results showed that running at all intensities produced significantly higher strains compared to walking (1985 ± 802 µε tensile, 5053 ± 181 µε compressive, p < 0.001), with peak tensile strains reaching 3731 µε and compressive strains up to 9541 µε. Low-intensity unilateral hopping also yielded significantly higher strains (3003 µε, p < 0.001) than walking, suggesting its osteogenic potential. In contrast, squat jumps, countermovement jumps, and bilateral hopping generated lower peak strains. The neural network model demonstrated high prediction accuracy, achieving correlations up to 0.97 and root mean square errors as low as 145.20 µε. These findings support the use of neural networks and IMU sensors for practical, cost-effective interventions to improve bone health and reduce fracture risk.

## 1. Introduction

Femoral neck fractures are the most common osteoporotic fractures, leading to high morbidity and mortality rates, with 50% of patients losing independent mobility and up to 30% mortality within six months^1,2^. Additionally, the related costs place a significant strain on healthcare systems. The number of men and women at high risk of experiencing a major osteoporotic fracture is projected to increase from 157 million in 2010 to 319 million by 2040^3^. Therefore, reducing the risk of osteoporotic hip fractures is critical.

Exercise is a proven method to enhance bone health. However, the type, intensity, and frequency of the exercise directly affect the extent of its benefits on bone health^4^. Ballistic exercises, such as hopping and jumping, are particularly promising for stimulating femoral neck adaptation^5,6^. It has been reported that normal walking^7^ is not associated with bone mineral density (BMD) changes in the femoral neck whereas jogging combined with walking^8^, running, and jumping^9,10^ were the most effective in improving BMD. On the other hand, while more frequent exercise is known to improve BMD, the specific benefits of increasing exercise intensity for enhancing BMD remain unclear^4^. Recent research has focused more on the effectiveness and safety of moderate-to high-intensity exercise compared to traditional low-intensity approaches, which prioritize safety^11,12^. A comprehensive meta-analysis examined the effects of different exercise intensities—low, moderate, and high—across various regimens, including resistance training, impact training such as walking, jogging, and jumping, and combined resistance and impact exercises on BMD at the lumber spine and femoral neck in postmenopausal women. The analysis revealed that high-intensity exercise was significantly more effective in increasing lumbar spine BMD compared to moderate– and low-intensity exercises, with mean differences of 0.031 g/cm^2^, 0.012 g/cm^2^, and 0.010 g/cm^2^, respectively^12^. However, at the femoral neck, low– and moderate-intensity exercise were equally effective, showing a mean difference of 0.011 g/cm^2^, while high-intensity exercise had no significant effect. In contrast, another study has showed that 6 months of unilateral, high-impact exercise of multidirectional hops completed daily increased the mean femoral neck BMD by 0.81% in postmenopausal women aged between 55 and 70 years^13^. Previous computational modeling studies have simulated the femoral neck response to various exercises, represented by mechanical strains, enabling the ranking of exercises based on their osteogenic potential^14^. The osteogenic response is triggered in areas where strain exceeds habitual loading levels, typically associated with normal walking^15,16^. A review by Martelli et al.^14^ reported that fast walking, but not necessarily running, optimally loads the femoral neck, while high-intensity jumps and hopping generate higher strains in the femoral neck than walking^15,17^. However, a previous study by the same author found that vertical and squat jumps produced lower femoral neck strains than walking, while one-leg long jumps resulted in higher strains^16^. These inconsistent findings highlight potential concerns regarding the impact of high-intensity exercises on joint health, raising safety considerations. Therefore, a comprehensive understanding of the mechanical response of the femoral neck to various ballistic exercises across different intensity levels is critical for designing effective preventive interventions for bone health.

Currently, no clinical method exists to directly measure the in-vivo mechanical response of the femoral neck for a certain type of locomotion. In biomechanical research, the “gold standard” non-invasive approach for predicting femoral neck response (strains) is musculoskeletal modeling (MSK) combined with finite element analysis (FE)^16–22^. The coupled MSK-FE model integrates two critical types of data: (1) three-dimensional (3D) bone architecture and density from medical imaging, such as computed tomography (CT) or magnetic resonance imaging (MRI), for the FE modeling component, and (2) muscle and joint contact forces, typically estimated through inverse dynamics and static optimization,based on 3D motion capture data, for the MSK modeling component. The predictions from MSK-FE models have the potential to significantly enhance fracture risk assessments, guide more effective treatment strategies, and improve rehabilitation protocols for clinicians, practitioners, and physiotherapists^23,24^. However, the MSK-FE method is resource-intensive, requiring specialized equipment, expertise, and considerable time, make it unsuitable for routine clinical use. Therefore, there is a critical need for a rapid, cost-effective, and user-friendly non-invasive method that can accurately predict in-vivo femoral neck strains during various locomotion modes (e.g., ballistic exercises).

Machine learning has become a leading technological trend in recent biomechanical research^25^. The extensive availability of large datasets from wearable sensors has driven significant advancements in estimating variables that traditionally required costly lab setups, such as ground reaction forces and other derived metrics. Machine learning has demonstrated its ability to accurately predict various kinetic and kinematic variables merely from wearable sensor measurements (e.g., Inertial measurement units (IMU))^26^, requiring less expert intervention and eliminating the need for expensive equipment. Multiple machine learning studies have estimated ground reaction force (GRF)^27,28^, joint moments^29– 34^, and internal joint forces^35,36^ during various locomotion tasks, using measures such as accelerations and gyroscopes that are (or can be) measured by IMU sensors. Wouda et al.^27^ used an artificial neural network to estimate vertical GRF during running using accelerations and lower limb joint angles. The network demonstrated a high correlation (>0.90) with the actual GRF time series. Guo et al.^28^ used Nonlinear System Identification (NARMAX) model from directly measured acceleration data without including joint kinematics to estimate vertical GRF during walking. Their model achieved a prediction error as low as 3.8% when compared to GRF data obtained from pressure insoles. Stetter et al.^35^ used a neural network with two hidden layers, similar to Wouda et al.^27^, to estimate knee joint forces during various exercises, including walking, running at different speeds, cutting maneuvers, one-leg jump, and counter-movement jump, using data from two IMU sensors. The results showed a good agreement between the estimated joint forces and those calculated through inverse dynamics for vertical and anterior-posterior knee forces with correlation coefficients ranging from 0.60 to 0.94, and 0.64 to 0.90 respectively (Stetter et al., 2019). Matijevich et al.^36^ estimated peak tibial force during running using various machine learning techniques, including neural networks and LASSO (Least Absolute Shrinkage and Selection Operator) regression. They converted lab-based data into signals that could be feasibly measured with IMU sensors and a pressure-sensing insole, achieving an estimation accuracy with a root mean-squared error (RMSE) of 0.25 ± 0.07 body weights and an absolute percent error between lab-based measured forces and machine learning estimated forces of 2.6 %. A very recent study by Haribaba and Basu^37^ evaluated various machine learning models, including neural networks, in conjunction with FE data to accelerate the prediction of the mechanical response (represented by strains) in the acetabulum of a healthy hip joint and periprosthetic bone in total hip joint replacement during walking gait. The study utilized different input features, such as bone condition, body weight, fin size, and loading conditions. A strong correlation was found between the predicted FE strains and those estimated by the neural network, with a coefficient of determination of 0.87 and RMSE of 0.04. These studies demonstrate the feasibility of neural networks to estimate the internal loadings of lower limb joint structures. In a recent study, we presented eXplainable convolutional neural network (XCM) to estimate lower limb joint moments, including hip joint, from data from four IMU sensors of various locomotion tasks^31^. Excellent agreement was found between the XCM estimated and the MSK inverse dynamics calculated hip joint moments with a correlation coefficient of 0.98.

This study has two primary aims. The first aim is to investigate the osteogenic response of the femoral neck to various ballistic exercises, including walking, running, countermovement jumps, squat jumps,unilateral hopping, and bilateral hopping, at three different intensity levels—low, moderate, and high— using MSK-FE modeling. We will rank the tested exercises based on the predicted peak first principal strain (tensile strain) and third principal strain (compressive strain) at the femoral neck and analyze the statistical differences in the predicted strains compared to normal walking at a self-selected speed. Exercises that produce significantly higher strains than walking are considered more effective for promoting bone health, assuming that osteogenic responses occur where exercise induced strain surpasses that of regular walking^14,15^. The second aim of the study is to develop a neural network using XCM architecture model capable of estimating femoral neck strains during various ballistic exercises using IMU sensors data represented by acceleration and gyroscope measurements. While previous studies have estimated joint kinematics and kinetics during various locomotion modes, no research to date has directly estimated the mechanical response of the bone in joints (bone strains) using a body-worn sensor setup. In previous studies, the number of IMU sensors and their measurement locations are often determined heuristically, and the impact underlying the selection of these parameter values on prediction accuracy has not been discussed yet^38^. Therefore, we further investigate the effect of using a reduced number of IMU sensors on model prediction accuracy. That will be done based on data from: (1) a comprehensive set of seven IMU sensors covering the entire lower body range of motion (trunk, left and right thigh, shank and foot), and (2) a reduced IMU sensor configuration of three sensors positioned around the region of interest only, the hip joint (trunk, and left and right thigh). The results of this study could help overcome current limitations in predicting femoral neck strains, which typically rely on expensive data and specialized expertise, and open new possibilities for using these predictions in clinical settings, potentially aiding in the design of effective preventive interventions, such as exercise regimes targeting bone health enhancement.

## 2. Material and methods

### 2.1. Participants and data collection and processing

Motion capture and musculoskeletal data of the current study has been used from our previously published study^39^ (first article of this series). In summary, a cohort of forty (20 males and 20 females) active participants were recruited with age range of 18 to 70 years old (mean ±SD: age of 40.3±13.1 years; height 1.71±0.08 m; and mass 68.44 ±11.67kg). All participants were healthy with no lower limb joint replacement or serious injury within the last year of the recruitment. Ethical approval was obtained from the University of Essex Faculty of Science & Health Ethics Subcommittee (ETH2021-1155). A written consent form was obtained from all participants before participating. Each participant attended one session in the biomechanics labs of the University of Essex, where three successful trials of walking, running, countermovement jump, squat jump, unilateral hopping, and bilateral hopping were collected at three different self-reported intensity levels (maximum, medium or intermediate, and minimum). Details of the characteristics of each exercise can be found in the supplementary materials (Table 2.SM.).

Figure 1 shows the overall workflow of the current study. For each participant, thirty-eight retro-reflective markers were attached to the lower body (twenty-two individual markers were attached to the left and right superior iliac spines, anterior superior iliac spines and posterior superior iliac spines, medial and lateral femoral condyles, medial and lateral malleoli, lateral and posterior aspects of the calcaneus, the first and fifth metatarsals while tracking clusters consisting of four markers were attached to the distal lateral aspect of the thigh and the shank). Marker trajectories were recorded using fourteen 3D motion capture cameras (Vicon. Ltd., Oxford, UK, 200 Hz, filtered at 18 Hz with a zero-lag second order low pass Butterworth). Ground reaction forces were collected using two-floor force plates (Kistler, Winterthur, Switzerland, 2000 Hz, filtered at 50 Hz with a zero-lag 2nd order low pass Butterworth) positioned side by side. To verify the musculoskeletal model’s muscle force predictions, five electromyography (EMG) sensors (Norixon, AZ., USA, 2000 Hz, high-pass filtered 30 Hz, 4th order Butterworth, rectified and low-pass filtered at 10 Hz) were attached unilaterally to the dominant side of each participant targeting five different muscles: gluteus maximus, gluteus medius, rectus femoris, biceps femoris, and soleus following SENIAM guidance. Details of the data can be found in Altai et al.^39^

**Figure 1.**
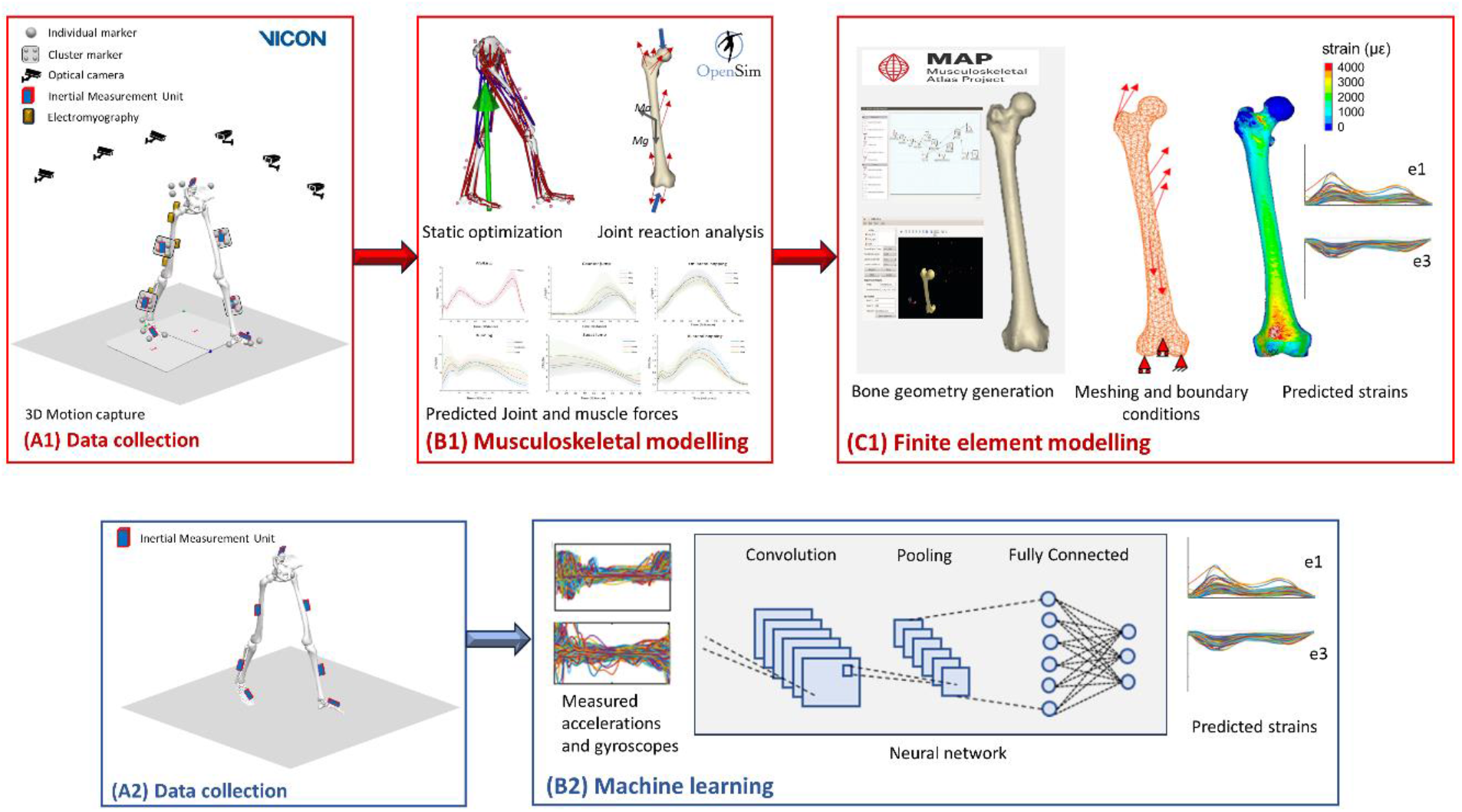
Framework followed in this study to predict the femoral neck strains using 1) typical musculoskeletal – finite element modelling pipeline (represented by A1, B1, and C1) and 2) the proposed neural network model (represented by A2 and B2). The typical modelling pipeline starts by collecting anatomical landmarks trajectories, ground reaction forces and electromyography signals in a 3D motion capture laboratory setting (A1), then musculoskeletal model is built using the collected data and generic model in OpenSim to estimated muscle and joint reaction forces using inverse dynamic and static optimization methods (B1), finally, finite element model is generated from three dimensional geometry of the femur and muscle and joint forces estimated by the musculoskeletal model to predict the femoral neck stains (first and third principal strains). The proposed pipeline predicts first and third principal stains merely from inertial measurement unit data (represented by accelerations and gyroscopes of the lower body segments) (A2) using neural network model (B2).

Seven Blue Trident IMU sensors (Vicon Motion Systems Ltd, Oxford, UK, 225Hz, filtered at 200 Hz with 2nd order low-pass Butterworth) were placed on the lower body segments of the left and right sides: posterior trunk, lateral shank, lateral thigh, and foot as shown in Figure 1 (A1). For each IMU sensor, accelerations and gyroscope data were recorded in the three planes of motion; sagittal, coronal (frontal), and transverse planes (represented by three axes x, y, and z), which were then used as the predictors for the neural network models.

All trials were processed in Vicon Nexus (V 2.12.1) and then the time of interest of each trail was segmented using Visual 3D (C-motion Inc., Germantown, MD, USA) as follows: walking and running from heal strike to toe-off of the same foot – a step, countermovement jump from the initial stand just before the take-off to the lowest position of the pelvis after landing, squat jump from the lowest position of the pelvis just before the take-off to the lowest position of the pelvis after landing, and for bilateral and unilateral hopping from the foot on to foot off the force plate of the same leg). The time of interest for all trials was defined using the dominant side of the participant. Data from all trials were then time normalized to 101 time points for musculoskeletal modelling.

### 2.2. Musculoskeletal models

A generic musculoskeletal model (gait2392)^40^ was modified by removing the torso and associated muscles (Figure 1 (B1)). Details of the musculoskeletal models can be found in Altai et al.^39^. In summary, the modified lower extremity model consisted of 13 body segments, 18 degrees of freedom (DOF), and 86 Hill-type musculotendon actuators. The hip was modelled as a ball and socket joint (3 DOF), while the knee was modeled as a sliding hinge joint (1 DOF rotational joint with translation coupled to the knee flexion angle), and the ankle and subtalar as revolute joints (1 DOF). Using OpenSim^41^, each model was scaled to match the subject’s anthropometric characteristics based on marker data of anatomical landmarks at the hip, knee and ankle during a static trial. Joint angles and moments were estimated using inverse kinematics and inverse dynamics, respectively, while static optimization was used to estimate muscle forces by minimizing the sum of squared muscle activations. Muscle attachment locations were extracted using a custom MATLAB (R2022b) script while force directions were determined using the muscle force direction plugin in OpenSim^42^. Muscle force directions together with muscle forces from static optimization were used to calculate muscle force components in x, y, and z. Hip, knee and ankle contact forces were calculated using joint reaction analysis^43^. The estimated muscle forces were then applied to the finite element models using the extracted muscle attachment location.

### 2.3. Finite element models

Since personalized medical images were not available, the three-dimensional geometry of the full femur for each participant was generated using the open-source Musculoskeletal Atlas Project (MAP) Client software^44^, which contains data from the Victorian Institute of Forensic Medicine (Melbourne, VIC, Australia). Using a generic lower-body shape model from the database, the full femur position and general size (surface mesh) were reconstructed from the anatomical landmarks of the participant’s motion capture data following the method described in Zhang et al.^44^. First, a generic whole lower body shape model was registered to the marker set defined in the static trial. Then, the atlas femur mesh was morphed into the femoral landmarks according to a femur statistical shape model^45^. The generated surface meshes of all participants were then imported into ANSYS (SpaceClaim 2023R1, PA, USA) to generate three-dimensional solid geometries, which were then meshed using 10-nodes tetrahedral elements in ANSYS (ICEM CFD 2023R1, PA, USA) with an average element size of 3mm^20,21^. Homogeneous linear elastic isotropic material properties were defined for the bone with an elastic modulus of 18.6 MPa and a 0.3 Poisson’s ratio^46^. Figure 1 (C1) summarize the steps for generating the finite element model.

Muscle forces were estimated by the musculoskeletal models and applied to the finite element model as point loads at the external surface of the femur. A list of included muscles can be found in Table 3.SM. in the supplementary materials. The location of the attachment points of each muscle was estimated by the musculoskeletal model and used to allocate the point of application of the force in the finite element model. Forces were then applied at the closest surface mesh node to the point of application estimated by the musculoskeletal model^20,21^. The distance between the point of application of the forces estimated by the musculoskeletal model and the closest nodes in the finite element model was less than the element size (3mm), except for three femurs, those were therefore excluded from the analysis leaving a cohort of 37 participants. The finite element models were kinematically constrained at the distal end of the femur to prevent rigid body motion ensuring that the equilibrium of the forces estimated by the musculoskeletal model was not disturbed. The most distal node of the medial condyle was fixed in all directions, while the displacement of the most distal node at the lateral condyle was constrained in the anterior-posterior and vertical (superior-inferior) directions. A third node in the patella groove was constrained anteroposteriorly^20,47^. These constraints were chosen to replicate the basic movements involved in the tested exercises, which are flexion-extension and rotation at the hip and knee joints; and abduction-adduction predominantly at the hip joint^48^.

Due to the high computational cost of the finite element models, only ten of the 101 timesteps were simulated for each trial. These time steps were carefully selected to include the peak point of the hip joint contact force curve as well as the first and last timesteps, ensuring full coverage of the trial period. At each of the ten timesteps, the peak first and third principal strains at the femoral neck were averaged across the surface nodes using a circle of 3mm radius, to follow the continuum hypothesis avoiding local effects of the load^49^. The location of the peak strains within the femoral neck region was also analyzed. All finite element simulations were performed in a local workstation using ANSYS Mechanical (APDL 2023R1, PA, USA). The computing time was on average one minute per timestep. The peak predicted strains at each of the ten times steps were then used as the outcomes in the neural network models.

### 2.4. Neural network models

First and third principal strains data predicted by the finite element models were used as the outcome dataset (2 variables represented by first and third principal strains) for the neural network model, while data of IMU sensors were the predictors (42 variables represented by accelerations and gyroscope of seven IMU sensors in three directions x, y, and z). Since neural networks are data hungry and to match time-series data of the predictors (101 timesteps), both first and third principal strains data were interpolated to regenerate 101 timesteps for each trial. Trials data of all exercises of all participants were combined for both the predictor and outcome datasets. The total number of observations in the dataset was 1729 corresponding to 1729 trials. The predictor dataset was organized into a 3D array shape 1729×42×101, where the second dimension was the number of predictors (accelerations and gyroscopes), and the third dimension was the number of time points. The outcome dataset was organized into a 2D array shape 1729×2×101, where the second dimension was the number of outcomes (first and third principal strains), and the third dimension was the number of time points. Each dataset was then split into training (75%, n = 1296) and testing (25%, n = 433) ensuring that the training and testing datasets were split with the same percentage for each exercise.

To assess the ability of the neural network to predict femoral neck strains for all tested exercises using a reduced number of IMU sensors, a subset of the predictor dataset was generated. This included data of three sensors (right thigh, left thigh, and trunk). The predictor subset data was also organized into a 3D array shape 1729×18×101 (18 variables represented by accelerations and gyroscope of three IMU sensors in three directions x, y, and z) while the outcome dataset was kept the same with 2D array shape 1729×2×101 (2 variables represented by first and third principal strains).

The neural network architecture was inspired by our previous work^31^ using eXplainable convolutional neural network XCM^50^. This neural network architecture has been shown to better predict biomechanical data from IMU sensors compared to other neural network architectures^31^. Figure 1 (2B) shows the XCM neural network model, the upper part of the XCM uses 2D convolution filters to extract features per observed variable and is composed of a 2D convolutional block, batch normalization, and ReLU activation layers. The lower part uses 1D convolution filters to extract information relative to time and captures the interaction between different time series. The output feature maps from these two parts are concatenated to form a feature map, which is passed through a 1D convolution block and global average pooling before performing classification with a softmax layer. The cyclical learning rate method was used to find the appropriate learning rate. The loss was plotted with respect to an increasing value of the learning rate. The learning rate was chosen to be in the interval that resulted in the lowest loss, which was found to be between 1e-1 and 2e-1. The learning rate took the value of 1e-1 at the first epoch and then gradually increased to reach a final value of 2e-1 at the last epoch (500 epoch). Analyses were performed in Python (version 3.9.0), with packages (Numpy v1.20.3, Pandas v1.3.4, Scipy v1.7.1) and models were trained using Tsai (version 0.3.1) from fastai with Google Collab.

### 2.5. Analysis

For the finite element predictions, initially, the curve of the peak femoral neck first and third principal strains in macrostrains along each trial period were found. Curves were then averaged across repetitive trials. The peak value of these averaged trials was then determined. The mean and standard deviation of the peak values for each exercise were then calculated and reported across all subjects. A repeated measure 2-way ANOVA was performed on the peaks of the first and third principal strains of all subjects to test the significant difference of femoral neck strains under various exercises compared to walking using the General Linear Model in SPSS (Chicago, USA). The dependent variable was the first and third principal strains, whilst the independent variables were the various exercise types and intensities. Where significance was found (significance level α = 0.05), Bonferroni post hoc test was conducted to quantify pairwise differences.

For the neural network predictions, for each exercise (including the three different levels), the agreement between the first and third principal strains estimated by the finite element models against their predicted values by the neural network model was derived from Pearson’s correlation coefficients (r)^51^, which were categorized as weak (r ≤ 0.35), moderate (0.35 < r ≤ 0.67), strong (0.67 < r ≤ 0.90) and excellent (r > 0.90). Additionally, the Root Mean Squared Error (RMSE) (Equ.1 SM. In supplementary materials), relative RMSE (relRMSE) (Equ.2 SM. In supplementary materials) expressed as a percentage (%) of the average peak-to-peak amplitude for the outcomes^52^ were determined to assess the accuracy of the neural network model predictions. RMSE, relRMSE, and r were assessed for each trial of each exercise and each participant, then means and standard deviations were then found for each exercise across all participants. The same analysis was conducted for: 1) the full set of seven IMU sensors and 2) the subset of three IMU sensors. Then relative difference between the two were evaluated to analysis the performance of the neural network with only three sensors compared to seven sensors.

## 3. Results

### 3.1. Strains predicted by finite element model

Exercises with various intensity levels were ranked with respect to the averaged peak first principal strain and averaged peak third principal strain (in macrostrains (×10^6^)) of the femoral neck predicted by MSK-FE as shown in Figure 2. Exercises with a significant difference (p < 0.05) compared to habitual walking at self-selected speed were marked with an asterisk. The estimates of the lower and upper limits and p-values as well as the results from a repeated measure 2-way ANOVA for General Linear Model were reported in supplemental material (Table 1.SM.). The mean and standard deviation of the average peak values for the first and third principal strains over the entire trial period for each exercise type and intensity level are reported in Table 1.

**Table 1.** Mean and standard deviation of the average peaks (throughout the entire trial period) of the vertical ground reaction force (vGRF) and hip joint reaction force (JCFhip), both normalized by the body weight, and of the peak first (e1) and third (e3) principal strains at the femoral neck predicted by the MSK-FE models for various exercise types and levels.

| Exercise | vGRF | JCFhip | e1 ($\mu\epsilon$) | e3 ($\mu\epsilon$) |
| --- | --- | --- | --- | --- |
| Walking | 1.28±0.09 | 6.31±1.23 | 1985 ± 802 | 5053 ± 181 |
| Running Fast | 2.51±0.26 | 8.21±1.32 | 3731 ± 1472 | 9541 ± 3610 |
| Running Moderate | 2.62±0.27 | 9.47±2.17 | 3534 ± 1197 | 9127 ± 2972 |
| Running Natural (slow) | 2.62±0.30 | 11.56±4.1 | 3389 ± 1321 | 8613 ± 2975 |
| Counter Movement Jumps Max | 1.05±0.15 | 4.06±1.33 | 1693 ± 1072 | 4190 ± 2470 |
| Counter Movement Jumps Med | 1.08±0.14 | 4.47±1.45 | 1392 ± 769 | 3377 ± 1742 |
| Counter Movement Jumps Min | 1.03±0.11 | 5.83±2.21 | 1164 ± 935 | 2715 ± 1148 |
| Squat Jumps Max | 1.22±0.18 | 4.18±1.52 | 1664 ± 935 | 3977 ± 2071 |
| Squat Jumps Med | 1.18±0.13 | 4.59±1.44 | 1302 ± 825 | 3003 ± 1310 |
| Squat Jumps Min | 1.19±0.14 | 5.63±2.38 | 1132 ± 744 | 2476 ± 1092 |
| Unilateral Hopping Max | 2.36±0.27 | 7.65±1.43 | 2439 ± 1061 | 6342 ± 2473 |
| Unilateral Hopping Med | 2.59±0.25 | 7.56±1.46 | 2729 ± 1112 | 7076 ± 2858 |
| Unilateral Hopping Min | 2.59±0.22 | 6.98±1.14 | 3003 ± 1189 | 7755 ± 3040 |
| Bilateral Hopping Max | 1.63±0.29 | 3.50±0.74 | 723 ± 407 | 1878 ± 926 |
| Bilateral Hopping Med | 1.76±0.24 | 3.18±0.57 | 795 ± 377 | 2050 ± 826 |
| Bilateral Hopping Min | 1.73±0.22 | 2.93±0.76 | 935 ± 459 | 2344 ± 1090 |

**Figure 2.**
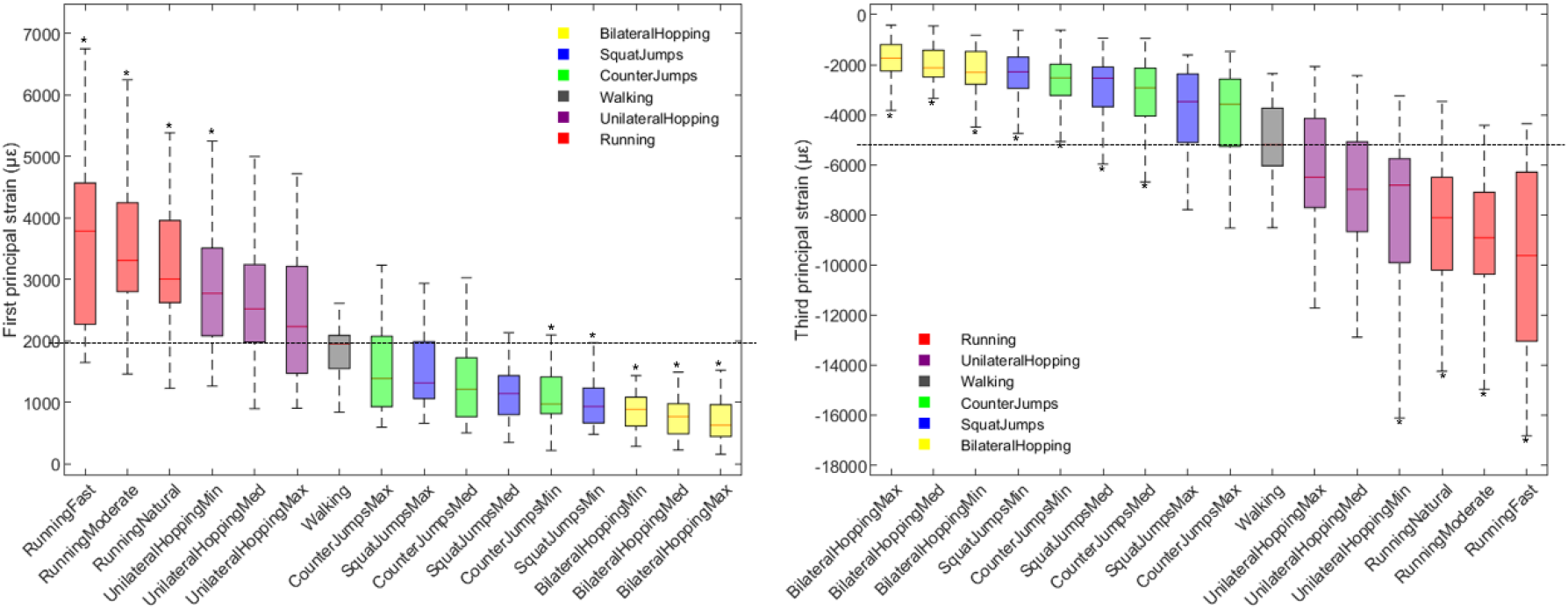
Box plot of large significant difference of peak first principal strain (left) and peak third principal strain (right) of the femoral neck under different levels of various ballistic exercises compared to walking at a self-selected speed indicated by the horizontal line. Asterisks denote the exercises with significant difference (*p < 0.05) compared to walking. Peak strains are ranked from left to right for the first principal strains and right to left for the third principal strains for the highest to the lowest estimated values for all included exercises.

Not all exercises demonstrated statistically significant differences in predicted peak strains compared to walking. Only running at all three intensity levels—fast speed 5.26 m/sec (3731 ± 1472 µε and 9541 ± 3610 µε for first and third principal strains, respectively), moderate speed 4.25 m/sec (3534 ± 1197 µε and 9127 ± 2972 µε), and natural speed 2.98 m/sec (3389 ± 1321 µε and 8613 ± 2975 µε)—and unilateral hopping at low intensity with a 0.31 sec stance duration (3003 ± 1189 µε) produced significantly higher peak strains (P < 0.001) than walking at a self-selected speed of 1.59 m/sec (1985 ± 802 µε and 5053 ± 181 µε for first and third principal strains, respectively). This indicates that, among the exercises tested, running at any intensity and low-intensity unilateral hopping have the potential to stimulate an osteogenic response in the femoral neck. In contrast, bilateral hopping at all intensity levels—maximum with stance duration of 0.19 sec (723 ± 407 µε and 1878 ± 926 µε), medium with stance duration of 0.21 sec (795 ± 377 µε and 2050 ± 826 µε), and minimum with stance duration of 0.25 sec (935 ± 459 µε and 2344 ± 1090 µε)—generated significantly lower peak strains than walking (P < 0.001). Similarly, the low-intensity squat jump with a 0.21 m jump height (1132 ± 744 µε and 2476 ± 1092 µε) and countermovement jump with a 0.23 m jump height (1164 ± 935 µε and 2715 ± 1148 µε) showed lower peak strains, with P-values of < 0.016 and < 0.028, respectively, for the first principal strain, and P < 0.001 for the third principal strain. While no significant differences were observed between walking and the squat jump at moderate and high intensities (0.28 m and 0.33 m jump heights, respectively) or the countermovement jump at moderate and high intensities (0.26 m and 0.32 m jump heights, respectively) (Table 1.SM). These findings suggest that neither of the two tested jump types nor bilateral hopping, at any intensity level, have the potential to induce an osteogenic response in the femoral neck.

The distribution of the first and third principal strains across the femoral head region under all tested exercises is shown in Figure 3 for a presentative case. Under walking and running, peak strains were located at the superior aspect of the femoral neck, noting that the second peak strains region where at the inferior aspect region. For both types of jumping and hopping exercises, peak strain locations shifted toward the inferior aspect of the femoral neck. Changing exercise intensity did not show any effect on the peak strain location at the femoral neck region.

**Figure 3.**
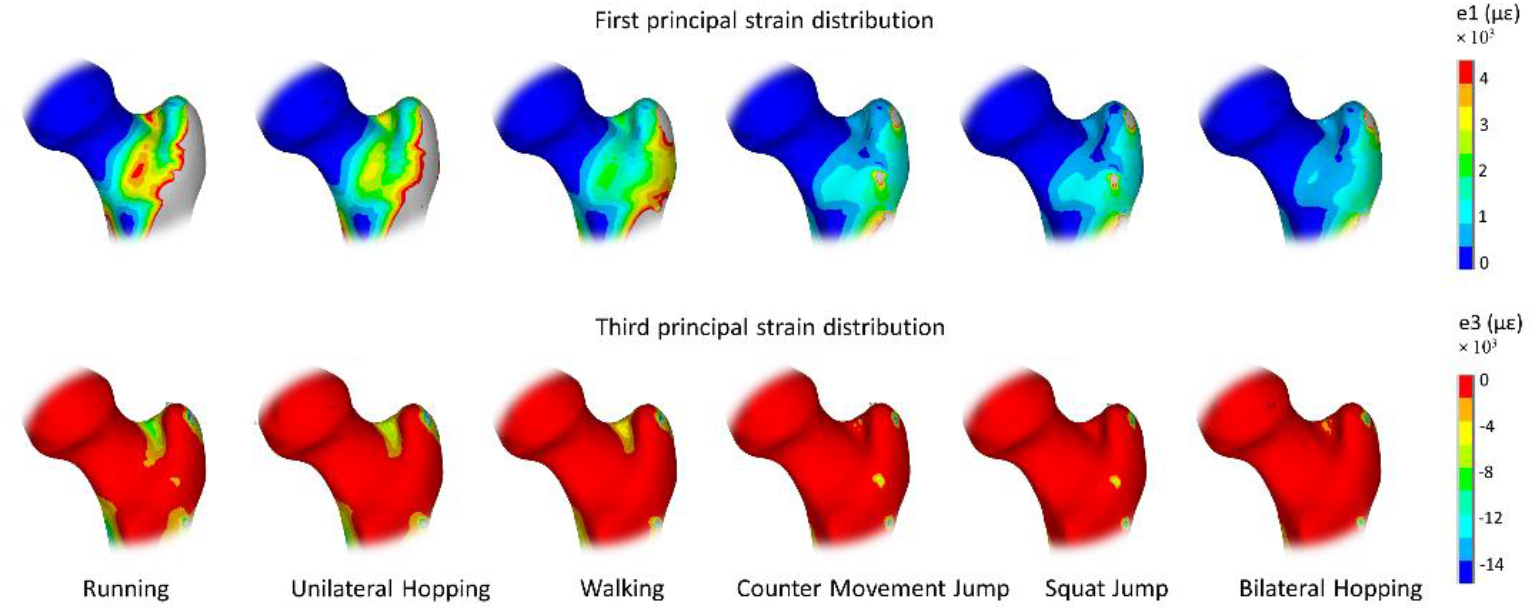
Distribution of the first and third principal strains predicted by MSK-FE model at the femoral neck under various ballistic exercises of a representative case.

### 3.2. Strains predicted by neural network model

Figure 4 illustrates the mean curves for the first and third principal strains, as estimated by the neural network model for all exercises, using a comprehensive set of seven IMU sensors, while Figure 5 shows the predicted strains using a reduced set of three IMU sensors. These are compared with predictions from the MSK-FE model. An overview of the neural network model’s estimated accuracy across all exercises is provided in Table 2. The relative differences in neural network model accuracy of the reduced set of IMU compared to the comprehensive set, are also reported for all exercises.

**Figure 4.**
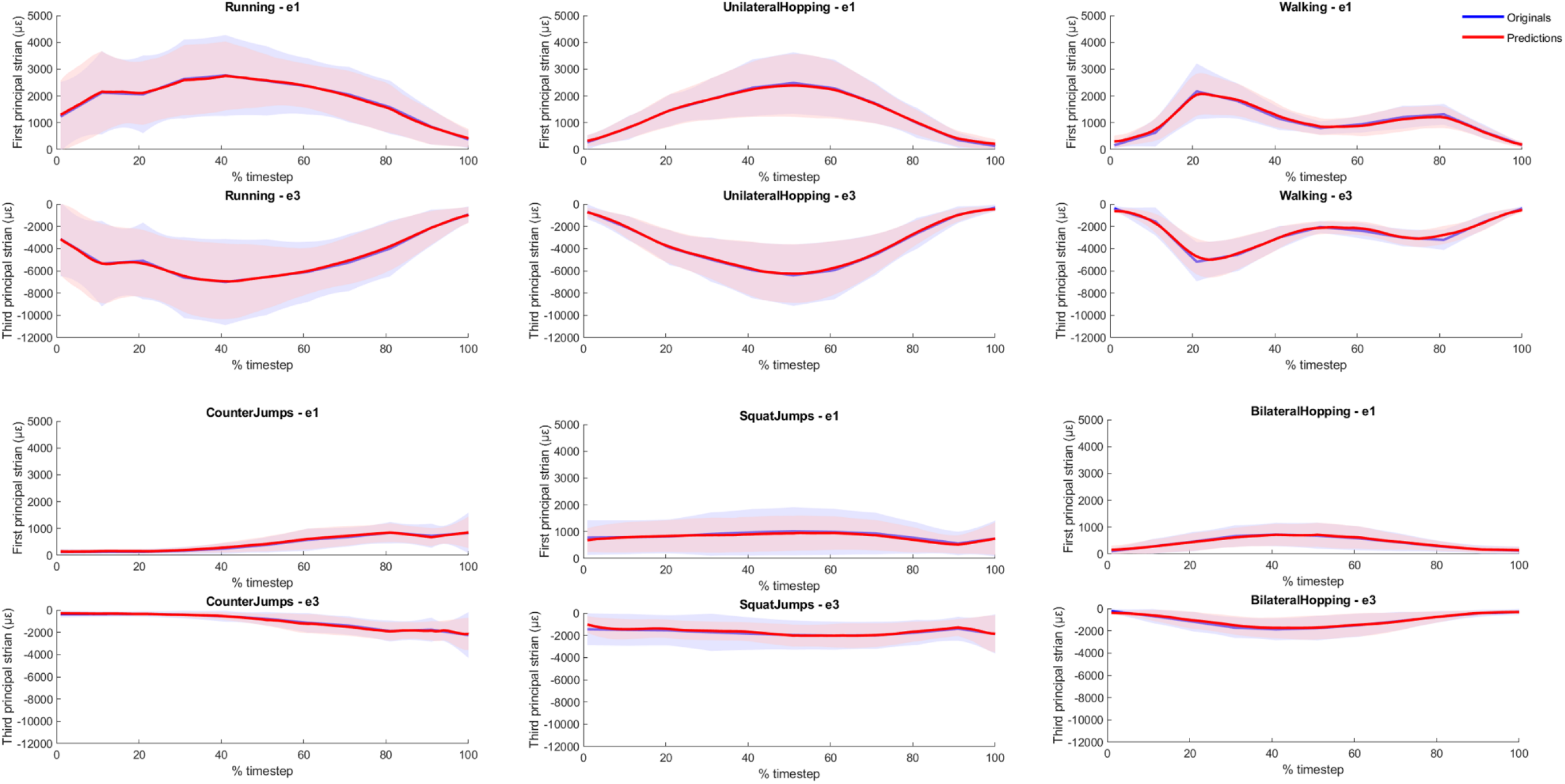
Mean first and third principal strains curves predicted by the XCM neural network compared to the original curves estimated by the finite element models for the different tested exercises using seven IMU sensors (trunk, right thigh, left thigh, right shank, left shank, right foot, and left foot).

**Figure 5.**
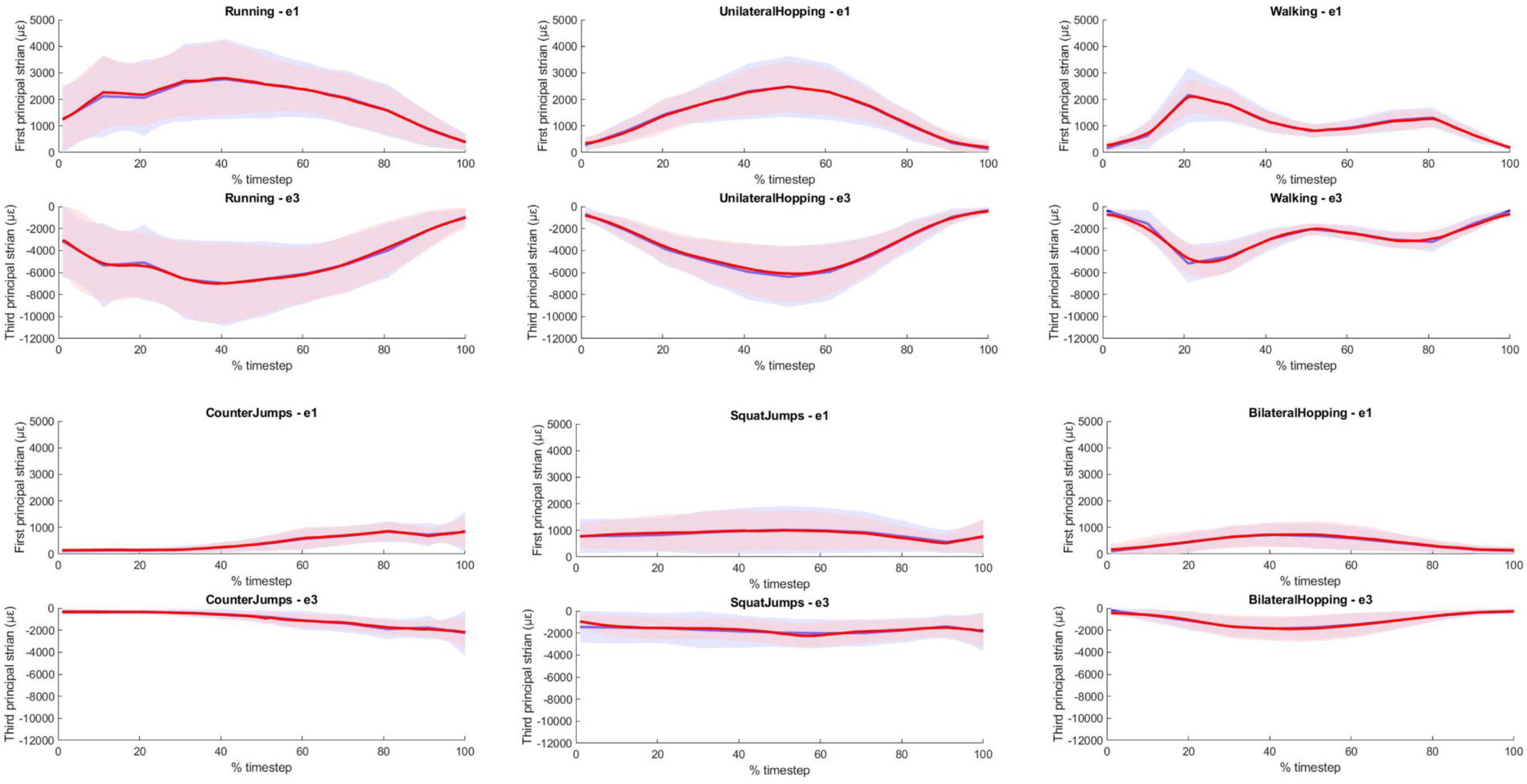
Mean first and third principal strains curves predicted by the XCM neural network compared to the original curves estimated by the finite element models for the different tested exercises using three IMU sensors (trunk, right thigh, and left thigh).

**Table 2.** Performance of XCM neural network in predicting first and third principal strains using two datasets: 1) data of seven IMU sensors, and 2) data of three IMU sensors, relative differences between the two also reported.

| Task | e1 ($\mu\epsilon$) | | | e3 ($\mu\epsilon$) | | |
| --- | --- | --- | --- | --- | --- | --- |
|  | RMSE | relRMSE (%) | r | RMSE | relRMSE (%) | r |
| **1) Using seven IMU sensors** |  |  |  |  |  |  |
| Walking | 182.85 ± 104.81 | 8.96 ± 3.80 | 0.96 ± 0.04 | 381.37 ± 119.22 | 8.01 ± 2.25 | 0.97 ± 0.02 |
| Running | 550.70 ± 412.97 | 17.86 ± 12.13 | 0.83 ± 0.27 | 1278.70 ± 797.85 | 16.51 ± 11.07 | 0.84 ± 0.26 |
| Unilateral Hopping | 309.18 ± 164.55 | 14.02 ± 9.74 | 0.95 ± 0.13 | 841.99 ± 578.01 | 14.22 ± 11.05 | 0.95 ± 0.08 |
| Bilateral Hopping | 169.15 ± 96.73 | 24.26 ± 15.06 | 0.81 ± 0.25 | 452.68 ± 255.16 | 25.36 ± 15.02 | 0.83 ± 0.20 |
| Counter Jumps | 145.20 ± 75.02 | 14.96 ± 11.34 | 0.93 ± 0.11 | 400.26 ± 233.87 | 16.37 ± 12.41 | 0.88 ± 0.22 |
| Squat Jumps | 238.84 ± 300.41 | 23.52 ± 16.64 | 0.80 ± 0.21 | 522.66 ± 719.28 | 22.64 ± 15.64 | 0.78 ± 0.26 |
| Mean | 278.70 ± 281.05 | 18.21 ± 13.58 | 0.87 ± 0.21 | 683.61 ± 641.99 | 18.22 ± 13.55 | 0.86 ± 0.22 |
| **2) Using three IMU sensors** |  |  |  |  |  |  |
| Walking | 220.48 ± 124.98 | 10.73 ± 4.11 | 0.95 ± 0.04 | 446.26 ± 110.66 | 9.78 ± 4.01 | 0.94 ± 0.07 |
| Running | 531.22 ± 363.79 | 17.08 ± 11.27 | 0.85 ± 0.22 | 1532.62 ± 1026.49 | 19.49 ± 12.18 | 0.81 ± 0.25 |
| Unilateral Hopping | 333.26 ± 240.89 | 14.05 ± 9.87 | 0.95 ± 0.10 | 898.75 ± 453.46 | 14.99 ± 7.88 | 0.93 ± 0.13 |
| Bilateral Hopping | 191.70 ± 120.74 | 25.94 ± 13.76 | 0.81 ± 0.21 | 507.65 ± 295.52 | 28.55 ± 15.81 | 0.78 ± 0.26 |
| Counter Jumps | 156.78 ± 94.28 | 15.47 ± 11.39 | 0.92 ± 0.17 | 437.26 ± 224.95 | 18.80 ± 14.24 | 0.87 ± 0.19 |
| Squat Jumps | 248.91 ± 298.53 | 23.43 ± 13.30 | 0.79 ± 0.18 | 623.66 ± 749.58 | 27.25 ± 18.78 | 0.72 ± 0.28 |
| Mean | 289.85 ± 274.46 | 18.55 ± 12.98 | 0.87 ± 0.19 | 783.52 ± 728.63 | 20.95 ± 14.91 | 0.83 ± 0.23 |
| **Relative difference (%)** |  |  |  |  |  |  |
| Walking | 20.58 | 19.67 | 0.32 | 17.02 | 22.13 | 2.28 |
| Running | 3.54 | 4.33 | 2.37 | 19.86 | 18.05 | 4.13 |
| Unilateral Hopping | 7.79 | 0.17 | 0.38 | 6.74 | 5.47 | 2.64 |
| Bilateral Hopping | 13.33 | 6.92 | 0.56 | 12.14 | 12.56 | 5.26 |
| Counter Jumps | 7.97 | 3.44 | 0.92 | 9.24 | 14.86 | 1.98 |
| Squat Jumps | 4.22 | 0.40 | 1.31 | 19.32 | 20.37 | 7.89 |
| Mean | 4.00 | 1.87 | 0.01 | 14.62 | 15.03 | 4.11 |

Data is presented as mean ± standard deviations. $\mu\epsilon$ is macrostrains. Relative difference is the percentage of the relative differences of the neural network predictions using data of the three IMU sensors in respect to using data of seven IMU sensors

The predicted strain curves by the neural network revealed strong to excellent correlations for both the first and third principal strains when using data from a comprehensive set of seven IMU sensors and when using data from a reduced set of three IMU sensors (Table 2). When using data from seven IMU sensors, the highest correlation for the first and third principal strain was observed for walking (r = 0.96 ± 0.04 and r = 0.97 ± 0.04, respectively) and for unilateral hopping (r = 0.95 ± 0.13 and r = 0.95 ± 0.08, respectively) with excellent correlations. The lowest correlation was for the squat jump, but still with good correlation for both first and third principal strains (r = 0.80 ± 0.21 and r = 0.78 ± 0.26, respectively). Across all exercises, the RMSE for first principal strains ranged between 145.20 ± 75.02 µε (counter movement jump) and 550.70 ± 412.97 µε (running), whereas for third principal strains, that was between 400.26 ± 233.87 µε (counter movement jump) and 1278.70 ± 797.85 µε (running). The relRMSE ranged between 8.96 ± 3.80% (walking) and 24.26 ± 15.06% (bilateral hopping) for the first principal strain and between 8.01 ± 2.25% (walking) and 25.36 ± 15.02% (bilateral hopping) for the third principal strain.

Similar trends were observed when using data of a reduced set of three IMU sensors with very small reduction in the neural network estimated accuracy in respect to using data of a comprehensive set of seven IMU sensors. The relative difference for the RMSE ranged from 4% (running and squat jumps) to 21% (walking) for the first principal strains and from 7% (unilateral hopping) to 20% (running) for the first principal strains. Details of the relative differences of all exercises can be found in Table 2.

## 4. Discussion

This study had two primary aims: first, to investigate the osteogenic response of the femoral neck to various ballistic exercises and intensity levels by predicting strains using the gold-standard MSK-FE modeling approach; and second, to investigate the ability of a neural network model to estimate the predicted strains only from body-worn IMU sensors, thereby bypassing the expensive and time-consuming MSK-FE modeling approach traditionally used in biomechanical research. Our results demonstrated that running at any speed (from slow jogging to fast sprinting), and unilateral hopping with longer stance durations, have the potential to stimulate an osteogenic response in the femoral neck. In contrast, jumping on both legs, regardless of intensity level, did not show such potential. While the neural network demonstrated excellent accuracy in predicting MSK-FE-derived strains based solely from IMU sensors data, highlighting its promising potential for clinical integration. By using femoral neck strains as an indicator of femoral neck health, this method could enhance fracture risk assessment and inform more targeted interventions, offering a practical and efficient alternative for routine clinical use.

Our MSK-FE predictions showed that not all ballistic exercises tested in this study induced significantly higher femoral neck strains compared to walking. Among the various exercises and intensity levels, fast running (5.26 m/sec), moderate running (4.25 m/sec), slow running (2.98 m/sec), and unilateral hopping with the longest stance duration (0.25 sec, categorized as a low intensity level) generated statistically higher femoral neck strains than walking at 1.59 m/sec (P < 0.001), indicating a potential osteogenic effect of the femoral neck. However, bilateral hopping at faster speeds, characterized by shorter stance durations (0.28 sec for moderate intensity and 0.31 sec for high intensity), did not show statistically significant differences compared to walking, yet still produced higher strain values. The reduced strain in faster hopping was associated with lower ground reaction forces and joint contact forces (Table 1) compared to slower hopping which may explain the reduction in the strain values. Our findings align with previous studies^15–17^. Similar trends were observed by Pellikaan et al.^15^ with peak femoral neck strains during unilateral hopping and various running speeds (1.95 m/sec to 2.5 m/sec) exceeding those during walking at 1.11 m/sec. However, they reported noticeably higher strain values during running (tensile strain 5412 µε at 2.5 m/sec) compared to our prediction (tensile strain 3389 µε at 2.98 m/sec), and during hopping (tensile strain 10373 µε vs. our prediction of 3003 µε). These discrepancies can be attributed to several factors, including modelling methodology and differences in participant demographics, which very likely played a role. Pellikaan et al.’s study^15^ involved postmenopausal women with an average age of 63, whereas our cohort consisted of highly active, younger males and females, with an average age of 40 ranging from 18 years to 70 years old. Anderson and Madigan^53^ showed that younger participants (aged 25 ± 4 years) exhibited 9% higher ground reaction forces and 18% higher hip contact forces, and 59% larger peak strains in early stance phase compared to older participants (aged 79 ± 5 years) walking at the same speed. In contrast to running and unilateral hopping, other exercises (countermovement jumps, squat jumps, and bilateral hopping) generated lower peak strains compared to walking. Bilateral hopping at all intensities, as well as both jump types at their minimum intensity levels, produced significantly lower strains than walking (P < 0.001), indicating that these exercises are not recommended for promoting an osteogenic effect in the femoral neck. Even at maximum intensity, jumping exercises still produced lower strains than walking with no statistical differences compared to walking. Our findings align with those of Martelli et al. ^16^, who also reported lower tensile strain peaks for vertical jumps (≈2500 µε in their study, compared to our range of 1164 µε to 1693 µε) and squat jumps (≈1800 µε in their study, compared to 1132 µε to 1664 µε in ours) when compared to walking (≈2700 µε). Kersh et al.^17^ also reported lower strain overall the femoral neck region than walking during landing on both feet from a light jump in place.

The higher strains observed during running, unilateral hopping, and walking, compared to countermovement, squat jumps, and bilateral hopping can be attributed to the nature of the exercises. When landing on both feet, impact forces are distributed across both legs, on the other hand, walking, running, and unilateral hopping involve periods where ground reaction forces act on a single limb. Consequently, our findings suggest that exercises involving bilateral jumps may be less effective in promoting femoral neck health compared to exercises involving unilateral jumps like running and unilateral hopping. This finding is also supported by several previous clinical trial studies, which indicated that fast walking programs^54,55^ and hopping exercises^56,57^ can lead to an increase in femoral neck BMD in elderly populations.

We observed that activities generating relatively similar ground reaction forces can produce relatively different levels and distributions of strain in the femoral neck. For instance, high-intensity unilateral hopping resulted in 34% higher peak strain compared to walking, and slow speed running (low intensity) produced 41% more strain than walking, despite both activities having roughly equivalent peak vertical ground reaction forces (Table 1). Forward propelling activities such as walking and running exerted maximal load on the superior region of the femoral neck, the thinnest part of the cortex, while, jumping in place shifted the peak strain towards the inferior aspect of the femoral neck (Figure 3), aligning with previous findings^15,17,20^. These results indicate that, when assessing femoral neck loading, the traditional assumption that the mechanical load correlates directly with ground reaction force^58^ requires reconsideration. The type of activity itself is a key determinant of femoral neck loads. We propose that the distinct anatomical arrangements and activation patterns of muscle groups around the hip contribute to varying mechanical stimuli on the femoral neck. For example, the gluteus maximus exerts direct effects on neighboring bone regions, while muscles like the semimembranosus influence hip-joint reaction forces indirectly. Additionally, muscles not spanning the hip may still contribute to these forces, albeit to a lesser extent, by dynamically accelerating body segments through musculoskeletal coupling^59,60^.

In general, agreement between neural network predictions and MSK-FE predictions ranged from excellent (r > 0.90 and relRMSE ≤ 15%) to good (r > 0.78 and relRMSE ≤ 25%) for all the ballistic exercises analyzed. Among these, walking demonstrated the highest estimation accuracies (r = 0.96 and relRMSE = 8.96%, r = 0.97 and relRMSE = 8.01% for first and third principal strains respectively), while there was a pronounced drop in estimation accuracies of the squat jump predictions (r = 0.80 and relRMSE = 23.52%, r = 0.78 and relRMSE = 22.64% for first and third principal strains respectively). One potential reason for the superior predictive power of walking is that it is performed at a consistent, self-selected intensity level, while other exercises are performed across three distinct intensity levels (maximum, medium, and minimum), which introduces a higher degree of variation in the movement dynamics. This variability could make it more challenging for the neural network model to generalize and predict accurately across all intensities. The increased variability in execution during squat jumps is further reflected in the high standard deviation for both the first and third principal strains across the cohort, indicating a wider dispersion in strain values among participants (Figure 4 and Figure 5). Similarly, running showed lower predictive accuracies (r = 0.83 and relRMSE = 17.86%, r = 0.84 and relRMSE = 16.07% for first and third principal strains respectively). The reduction in accuracy for running can also be attributed to the high inter-individual variability in predicted strain values, which is a common characteristic of more dynamic and explosive movements. This trend between predictive accuracy reduction and higher rate of data variation is consistent with findings reported by Setter et al.^35^ and Fluit et al.^61^. Stetter et al.^35^ observed a similar reduction in the accuracy of knee joint force predictions made by a neural network model for walking, which was associated with higher variability in knee joint forces, compared to running. Similarly, Fluit et al.^61^ observed similar changes in estimation accuracy when they assessed a prediction model for ground reaction forces and moments during various daily activities using 3D full-body motion analysis. This suggests that the performance of a machine learning model is sensitive to the consistency of movement and the level of variability in the data it is trained on. Furthermore, this may indicate that the model should not be trained on generalized data; instead, it should be population specific if precise accuracy is required. However, the reduction in model accuracy observed in our study was minimal. Future studies should further investigate the impact of data variability on machine learning prediction accuracy.

Distinct differences in neural network estimation accuracy were seen between unilateral hopping and bilateral hopping (r = 0.96 and relRMSE = 14.02 %, r = 0.81 and relRMSE = 24.26% respectively). This has also been observed in Setter et al. study^35^, where model accuracy for predicting knee joint forces was lower for two-leg jumps than for one-leg jumps (r for take-off = 0.92 vs. 0.60; r for landing = 0.84 vs. 0.61). One reason for the reduced estimation accuracy for bilateral hopping may be the bipedal characteristic of the movement. Potential inaccuracies in the strain estimations are caused by the distribution of the total external load on both legs. Stetter et al.^35^ suggested that incorporating an activity recognition approach could help mitigate these limitations. By selecting individualized prediction models based on specific movement categories, the model could account for the distinct characteristics of different movement types and improve accuracy.

Reducing the training data for the neural network model by using only three IMU sensors instead of seven had minimal impact on prediction accuracy, which remained excellent (r = 0.95, relRMSE < 16%) to good (r = 0.72, relRMSE < 16%) across all tested exercises, following similar trends as with seven IMUs. Walking showed the largest reduction in prediction accuracy, with relative differences compared to the seven-sensor setup as r = 0.32%, relRMSE = 19.62%, and RMSE = 20.58% for first principal strains, and r = 2.28%, relRMSE = 22.13%, and RMSE = 17.02% for third principal strains. This again may be due to the imbalance in the data size between walking (fewer trial numbers) and other exercises, especially, when using minimal training data size. This imbalance could lead to a neural network bias toward other exercises, reducing walking prediction accuracy. Type and size of training data plays a key role in improving neural network training efficiency and test accuracy^62^. Additionally, by excluding sensors from the shank and foot, important biomechanical data—such as foot strike patterns, ankle movements, and shank rotation—are not captured, limiting the model’s ability to fully understand lower limb mechanics and so struggle to capture important biomechanical details. However, in general, the reduction in accuracy of our neural network was minimal, and the performance remained nearly as high as when using seven IMU sensors. Our finding is supported by a number of recent studies which used artificial neural network with a limited amount of IMU measurement information, but to predict ground reaction forces during walking and running^28,38,63^. For example, Guo et al.^28^ used a single IMU measurements taken at the sacrum to predicted the vertical ground reaction forces and reported an average prediction error of less than 5.0% for walking. Ngoh et al.^63^ demonstrated that neural network predict vertical ground reaction forces with one uniaxial IMU sensor located at the foot with average errors ranging between the 0.10 and 0.18 of body weight at different running speeds. It may be important to note that a full set of IMUs may be beneficial where high precision is required, depending on the application. However, if the goal is to obtain an indication of strain patterns and levels during a specific exercise, our results suggest that a reduced set of IMU sensors can still provide sufficient accuracy. This may be especially advantageous when considering cost, data size, and time.

One of the main limitations of the current study is that strain data predicted by the MSK-FE model were not fully personalized. Multi-level personalization of neuromusculoskeletal models can significantly influence the estimation of internal loading^64^. As a result, our MSK-FE predictions may not entirely capture the individual variations among participants, which could also impact the accuracy of the neural network predictions^62^. This limitation was primarily due to the lack of available data, particularly the absence of medical images necessary to create personalized FE models for each participant. A full set of personalized data, required of such modelling pipeline, has been and still is a big challenge in the biomechanical field. Previous studies have relied on body-matched volunteers^16,65,66^, synthetic model^19^, or generic scaled bone model^67^, similar to the approach followed in our study. In the future, our model can be tested with personalized data as it becomes available. Another limitation is that hyperparameter tuning has not been explored in the current study, hence, our findings can provide a more conservative estimation of the predictive performance of the neural network. Lastly, the characteristics of the participants (e.g., sex and age group) may be important determinants in model prediction accuracy. A machine-learned model used for prediction purposes must be trained on data that has similar characteristics to the data needed to be predicted. Yet, although our cohort included a large age group ranging from 18 to 70 years, all participants were healthy and active individual who exercise regularly which was confirmed by the relatively lower variation in the estimated joint forces and strains.

In this work, the increased strain observed in activities like running and unilateral hopping, compared to walking, suggests these exercises could form the basis for early intervention strategies aimed at enhancing bone health and mitigating fracture risk over the course of a lifetime, particularly in individuals at risk of osteoporosis. These findings emphasize the potential benefits of incorporating high-impact, weight-bearing exercises into preventive and therapeutic programs to stimulate bone adaptation and improve skeletal strength. Additionally, we demonstrate that combining IMU sensor data with neural network modeling provides an efficient and accurate method for predicting femoral neck strain during ballistic exercises. This approach is significantly faster than traditional MSK-FE modeling, which can take hours or days due to complex processes like model scaling, 3D reconstruction, and meshing. This makes it an accessible, scalable tool for both clinical and sports applications, reducing reliance on specialized expertise and high-end computational resources. Moreover, this method opens the possibility for near-real-time biomechanical analysis, facilitating timely and practical insights into bone health and injury prevention in various settings.

## 5. Author Contributions

Conceptualization—ZA, BL; Data Curation-ZA, Formal Analysis—ZA, BL, Funding Acquisition— BL, AP, JM Methodology—ZA, BL, Project Administration—BL, Software—ZA, Supervision—AP, JM, BL, Validation—AP, JM, QM, BL, Visualization—ZA, Writing—Original Draft Preparation— ZA, Writing—Review and Editing-All authors.

## 7. Acknowledgments

This work was supported by The Academy of Medical Sciences, UK, Springboard Award (SBF006\1019).

## 8. Supplementary Material

The Supplementary Material for this article can be found online with the article.

## 9. Data Availability Statement

All data produced in the present study are available upon reasonable request to the authors.

