## Supplementary Materials for "From Exercise to Strain: Rapid and Accurate Prediction of Femoral Neck Loading"

*** Correspondence:**
Zainab Altai

Colchester, Essex, UK

+44 (0) 1206 876217 


**Table 1.SM.** The estimates, lower/upper limits, and p-values as well as the results from the ANOVA test on the peak first and third principal strain at the femoral neck. Exercises with a significant difference (p < .05) compared to walking are marked with an asterisk.

|  | **Exercise** | **Mean Difference (walking-exercise)** | **P value** | **95% Confidence Interval for Difference** | |
| --- | --- | --- | --- | --- | --- |
|  |  |  |  | **Lower Bound** | **Upper Bound** |
| First principal strain | RunningNatural | -.001404* | <.001 | -.002191 | -.000616 |
|  | RunningModerate | -.001549* | <.001 | -.002337 | -.000761 |
|  | RunningFast | -.001746* | <.001 | -.002534 | -.000958 |
|  | SquatJumpsMin | .000853* | .016 | .000066 | .001641 |
|  | SquatJumpsMed | .000683 | .261 | -.000105 | .001471 |
|  | SquatJumpsMax | .000321 | 1.000 | -.000466 | .001109 |
|  | CounterJumpsMin | .000821* | .028 | .000034 | .001609 |
|  | CounterJumpsMed | .000593 | .928 | -.000195 | .001381 |
|  | CounterJumpsMax | .000293 | 1.000 | -.000495 | .001080 |
|  | UnilateralHoppingMin | -.001018* | <.001 | -.001806 | -.000230 |
|  | UnilateralHoppingMed | -.000744 | .102 | -.001532 | .000043 |
|  | UnilateralHoppingMax | -.000454 | 1.000 | -.001242 | .000333 |
|  | BilateralHoppingMin | .001050* | <.001 | .000262 | .001838 |
|  | BilateralHoppingMed | .001190* | <.001 | .000403 | .001978 |
|  | BilateralHoppingMax | .001263* | <.001 | .000475 | .002050 |
| Third principal strain | RunningNatural | .003560* | <.001 | .001751 | .005368 |
|  | RunningModerate | .004074* | <.001 | .002265 | .005882 |
|  | RunningFast | .004495* | <.001 | .002687 | .006304 |
|  | SquatJumpsMin | -.002577* | <.001 | -.004386 | -.000769 |
|  | SquatJumpsMed | -.002051* | .008 | -.003859 | -.000242 |
|  | SquatJumpsMax | -.001077 | 1.000 | -.002885 | .000732 |
|  | CounterJumpsMin | -.002339* | <.001 | -.004147 | -.000530 |
|  | CounterJumpsMed | -.001676 | .127 | -.003485 | .000132 |
|  | CounterJumpsMax | -.000864 | 1.000 | -.002672 | .000945 |
|  | UnilateralHoppingMin | .002701* | <.001 | .000893 | .004510 |
|  | UnilateralHoppingMed | .002022* | .010 | .000214 | .003831 |
|  | UnilateralHoppingMax | .001288 | 1.000 | -.000520 | .003097 |
|  | BilateralHoppingMin | -.002709* | <.001 | -.004518 | -.000901 |
|  | BilateralHoppingMed | -.003004* | <.001 | -.004812 | -.001195 |
|  | BilateralHoppingMax | -.003176* | <.001 | -.004984 | -.001367 |

**Table 2.SM.** Mean and SD of the Walking ad running speed, jump height, and stance duration for hopping for the intensity level of each exercise.

| **Exercise** | **Mean±SD Speed (m/sec)** | **Mean±SD Jump height (m)** | **Mean±SD Stance duration (sec)** |
| --- | --- | --- | --- |
| Walking | 1.59±0.41 | - | - |
| Running Natural | 2.98±0.61 | - | - |
| Running Moderate | 4.25±0.59 | - | - |
| Running Fast | 5.26±0.83 | - | - |
| Squat Jumps Min | - | 0.21±0.06 | - |
| Squat Jumps Med | - | 0.26±0.08 | - |
| Squat Jumps Max | - | 0.32±0.09 | - |
| Counter Jumps Min | - | 0.23±0.05 | - |
| Counter Jumps Med | - | 0.28±0.07 | - |
| Counter Jumps Max | - | 0.33±0.08 | - |
| Unilateral Hopping Min | - | - | 0.31±0.05 |
| Unilateral Hopping Med | - | - | 0.28±0.06 |
| Unilateral Hopping Max | - | - | 0.25±0.06 |
| Bilateral Hopping Min | - | - | 0.25±0.04 |
| Bilateral Hopping Med | - | - | 0.21±0.03 |
| Bilateral Hopping Max | - | - | 0.19±0.02 |

**Table 3.SM.** List of muscles included in the finite element model and were estimated by the musculoskeletal model.

**Muscle**

Adductor Brevis

Adductor Longis

Adductor Magnus

Biceps Femoris long head

Biceps Femoris short head

Gemellus

Gluteus Maximus

Gluteus Medius

Gluteus Minimus

Iliacus

Gastrocnemius Lateralis

Gastrocnemius Medialis

Pectineus

Piriformis

Psoas

Quadratus Femoris

Vastus Intermedius

Vastus Lateralis

Vastus Medialis

$RMSE=\sqrt{\frac{\int_{0}^{T} \left[ u_{obs}\left( t \right)-u_{pred}(t) \right]^{2}dt}{T}}$ (**Equ1. SM.**)

$relRMSE=\frac{\mathrm{RMSE}}{0.5\left[ \sum_{i=1}^{2} \left( {max}_{0<t<T}\left( u_{i}\left( t \right) \right)-{min}_{0<t<T}\left( u_{i}\left( t \right) \right) \right) \right]}\times100\%$ (**Equ2. SM.**)

where *T* is trial period, $u_{obs}\left( t \right)$ is the value at the $t^{th}$ time point of the observed outcome, $u_{pred}(t)$ is the value at the $t^{th}$ time point of the predicted outcome, and *i* represents either the observed or predicted outcomes. Root mean squared error (RMSE), relative RMSE (relRMSE) expressed as a percentage (%) of the average peak-to-peak amplitude for the outcomes.
